# REACT-1 round 13 interim report: acceleration of SARS-CoV-2 Delta epidemic in the community in England during late June and early July 2021

**DOI:** 10.1101/2021.07.08.21260185

**Authors:** Steven Riley, Oliver Eales, David Haw, Haowei Wang, Caroline E. Walters, Kylie E. C. Ainslie, Christina Atchison, Claudio Fronterre, Peter J. Diggle, Deborah Ashby, Christl A. Donnelly, Wendy Barclay, Graham Cooke, Helen Ward, Ara Darzi, Paul Elliott

## Abstract

**Background:** Despite high levels of vaccination in the adult population, cases of COVID-19 have risen exponentially in England since the start of May 2021 driven by the Delta variant. However, with far fewer hospitalisations and deaths per case during the recent growth in cases compared with 2020, it is intended that all remaining social distancing legislation in England will be removed from 19 July 2021.

**Methods:** We report interim results from round 13 of the REal-time Assessment of Community Transmission-1 (REACT-1) study in which a cross-sectional sample of the population of England was asked to provide a throat and nose swab for RT-PCR and to answer a questionnaire. Data collection for this report (round 13 interim) was from 24 June to 5 July 2021.

**Results:** In round 13 interim, we found 237 positives from 47,729 swabs giving a weighted prevalence of 0.59% (0.51%, 0.68%) which was approximately four-fold higher compared with round 12 at 0.15% (0.12%, 0.18%). This resulted from continued exponential growth in prevalence with an average doubling time of 15 (13, 17) days between round 12 and round 13. However, during the recent period of round 13 interim only, we observed a shorter doubling time of 6.1 (4.0, 12) days with a corresponding R number of 1.87 (1.40, 2.45). There were substantial increases in all age groups under the age of 75 years, and especially at younger ages, with the highest prevalence in 13 to 17 year olds at 1.33% (0.97%, 1.82%) and in 18 to 24 years olds at 1.40% (0.89%, 2.18%). Infections have increased in all regions with the largest increase in London where prevalence increased more than eight-fold from 0.13% (0.08%, 0.20%) in round 12 to 1.08% (0.79%, 1.47%) in round 13 interim. Overall, prevalence was over 3 times higher in the unvaccinated compared with those reporting two doses of vaccine in both round 12 and round 13 interim, although there was a similar proportional increase in prevalence in vaccinated and unvaccinated individuals between the two rounds.

**Discussion:** We are entering a critical period with a number of important competing processes: continued vaccination rollout to the whole adult population in England, increased natural immunity through infection, reduced social mixing of children during school holidays, increased proportion of mixing occurring outdoors during summer, the intended full opening of hospitality and entertainment and cessation of mandated social distancing and mask wearing. Surveillance programmes are essential during this next phase of the epidemic to provide clear evidence to the government and the public on the levels and trends in prevalence of infections and their relationship to vaccine coverage, hospitalisations, deaths and Long COVID.

## Introduction

The COVID-19 vaccination programme in the UK has been highly effective. As of 5 July 2021, 86% of people in England aged 18 or older had received one dose of vaccine against SARS-CoV-2 with 64% receiving two doses [1]. Both vaccines are effective in reducing the risk of hospitalisation and death [2] and have largely retained that efficacy [3] against the Delta variant, especially for those receiving two doses [4]. However, the Delta variant successfully replaced the previously dominant Alpha variant in England and established a trend of exponential growth at the beginning of June 2021 [5], resulting in a delay to the final stage (step four) of the roadmap for exiting the third national lockdown. The lockdown was implemented in early January 2021 to reduce pressure on healthcare services that were at risk of being overwhelmed [6]. Exiting the lockdown had been due to occur on 21 June 2021 but was delayed to allow the potential impact of Delta variant on hospitalisations and deaths to be better assessed [7].

Since step four of the roadmap for exiting lockdown was delayed, PCR-confirmed cases documented through routine testing (Pillar 1 and Pillar 2) have continued to increase in England. However, hospitalisations have increased far more slowly than during a period of rapid exponential growth during September and October 2020 [1], reflecting a partial ‘uncoupling’ of infections and hospitalisations associated with the successful roll-out of the vaccination programme [5]. The final stage of the roadmap for exiting lockdown is now scheduled to take place on 19 July 2021 with final confirmation on 12 July, under the assumption that eventual vaccine uptake and effectiveness will be high enough -- and the peak of infections low enough -- that pressure propagated through to healthcare services will be manageable [8].

Here we report interim results from round 13 of the REal-time Assessment of Community Transmission-1 (REACT-1) study in which throat and nose swabs are obtained from a representative sample of people in England aged 5 years and older [9,10]. Round 13 commenced on 24 June 2021 and we report here the results from swabs collected up to and including 5 July 2021 (round 13 interim). We compare these with complete results for round 12, in which swabs were collected from 20 May to 7 June 2021 [5], with the objectives of measuring the rate of change of the epidemic in England and identifying key drivers of that change (growth or decline).

## Results

We found 237 positives from 47,729 swabs giving a weighted prevalence of 0.59% (0.51%, 0.68%) (Table 1, Figure 1). This represents an approximately four-fold increase over weighted prevalence in round 12 of 0.15% (0.12%, 0.18%). Although average prevalence for round 13 interim is similar to that observed during round 5 (18 September 2020 to 5 October 2020), the rate of increase appears steeper.

**Table 1.** The unweighted and weighted prevalence of swab-positivity across 12 complete rounds of REACT-1 and round 13 interim.

| Round | Tested swabs | Positive swabs | Unweighted prevalence (95% CI) *** | Weighted prevalence (95% CI) | First sample | Last sample |
| --- | --- | --- | --- | --- | --- | --- |
| 1 | 120,620 | 159 | 0.13% (0.11%, 0.15%) | 0.16% (0.13%, 0.19%) | 1/5/2020 | 1/6/2020 |
| 2 | 159,199 | 123 | 0.077% (0.065%, 0.092%) | 0.088% (0.068%, 0.11%) | 19/6/2020 | 7/7/2020 |
| 3 | 162,821 | 54 | 0.033% (0.025%, 0.043%) | 0.040% (0.027%, 0.053%) | 24/7/2020 | 11/8/2020 |
| 4 | 154,325 | 137 | 0.089% (0.075%, 0.11%) | 0.13% (0.096%, 0.15%) | 20/8/2020 | 8/9/2020 |
| 5 | 174,949 | 824 | 0.47% (0.44%, 0.50%) | 0.60% (0.55%, 0.71%) | 18/9/2020 | 5/10/2020 |
| 6 | 160,175 | 1,732 | 1.08% (1.03%, 1.13%) | 1.30% (1.21%, 1.39%) | 16/10/2020 | 2/11/2020 |
| 7 | 168,181 | 1,299 | 0.77% (0.73%, 0.82%) | 0.94% (0.87%, 1.01%) | 13/11/2020 | 3/12/2020 |
| 8 | 167,642 | 2,282 | 1.36% (1.31%, 1.42%) | 1.57% (1.49%, 1.66%) | 06/01/2021* | 22/01/2021 |
| 9 | 165,456 | 689 | 0.42% (0.39%, 0.45%) | 0.49% (0.44%, 0.55%) | 4/2/2021 | 23/2/2021 |
| 10 | 140,844 | 227 | 0.16% (0.14%, 0.18%) | 0.20% (0.17%, 0.23%) | 11/03/2021 | 30/3/2021 |
| 11 | 127,408 | 115 | 0.09% (0.07%, 0.11%) | 0.10% (0.08%, 0.13%) | 15/04/2021 | 3/5/2021 |
| 12 | 108,911 | 135 | 0.12% (0.10%, 0.15%) | 0.15% (0.12%, 0.18%) | 20/05/2021 | 07/06/2021 |
| 13 interim | 47,729 | 237 | 0.50% (0.44%, 0.56%) | 0.59% (0.51%, 0.68%) | 24/06/2021 | 05/07/2021** |

\* Includes small number of samples from previous days
\*\* Not yet complete
\*\*\* Sampling strategy changed for round 12 and subsequent rounds. Therefore unweighted prevalence is not directly comparable with previous rounds

**Figure 1.**
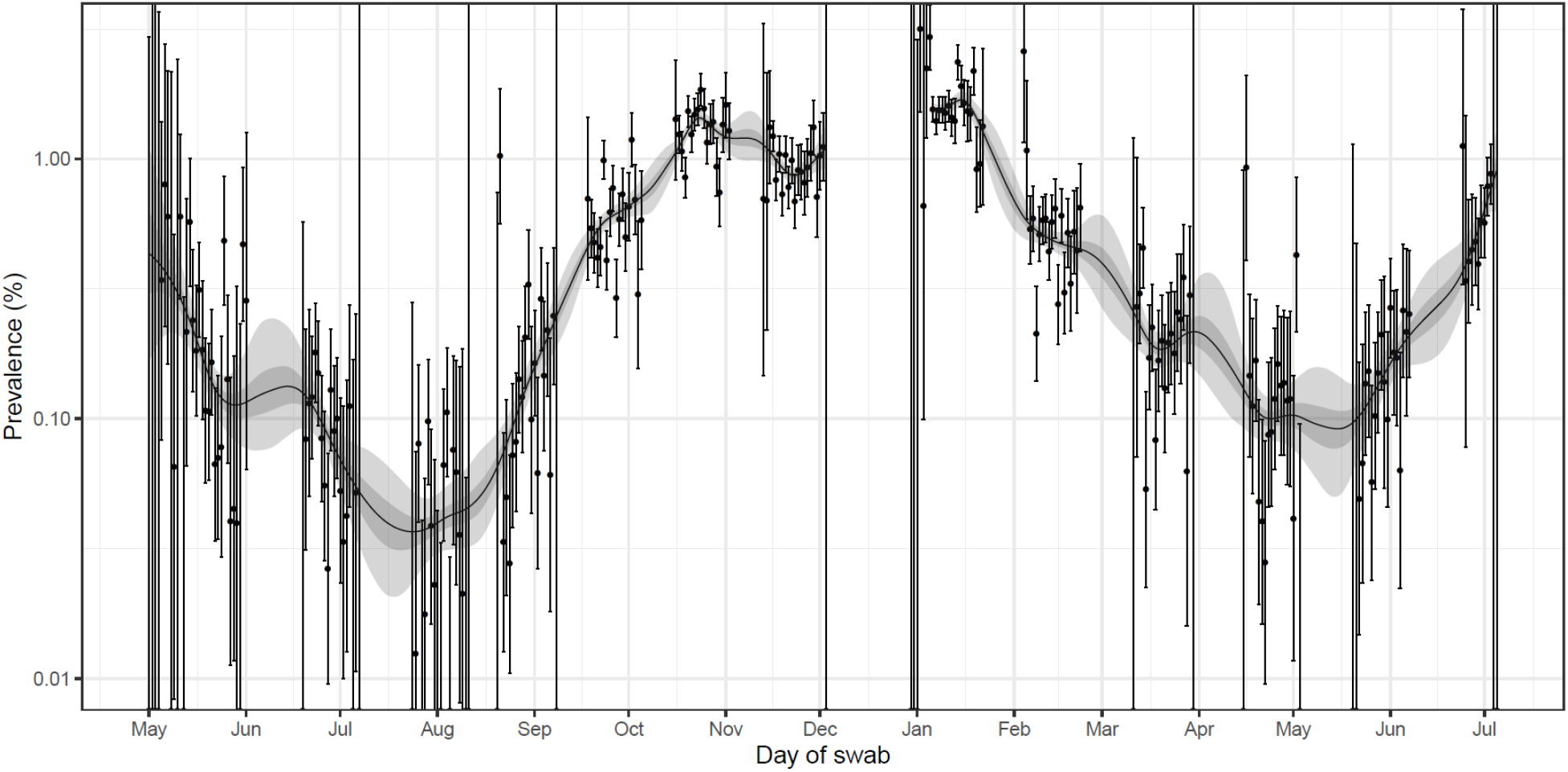
Prevalence of national swab-positivity for England estimated using a P-spline for all twelve completed rounds and round 13 interim with central 50% (dark grey) and 95% (light grey) posterior credible intervals. Shown here for the entire period of the study with a log10 y-axis. Weighted observations (black dots) and 95% binomial confidence intervals (vertical lines) are also shown. Note that the period between round 7 and round 8 (December) of the model is not included as there were no data available to capture the late December peak of the epidemic.

Using constant growth rate models, we found evidence for continued exponential growth between round 12 and round 13 interim with a doubling time of 15 (13, 17) days (Table 2, Figure 2). However, we also observed an acceleration during round 13 interim for which we estimated a doubling time of 6.1 (4.0, 12) days and a corresponding R of 1.87 (1.40, 2.45). An acceleration was also apparent as an upward curve in the P-spline model of prevalence (Figure 1).

**Table 2.** Estimates of national growth rates, doubling times and reproduction numbers for round 13 interim, and round 12 to round 13 interim.

| Round | Outcome | Growth rate | R | Probability R>1 | Doubling (+) / Halving (-) time |
| --- | --- | --- | --- | --- | --- |
| 12 and 13 interim | All positives | 0.046 ( 0.040 , 0.052 ) | 1.32 ( 1.27 , 1.37 ) | >0.99 | 15.1 ( 17.4 , 13.2 ) |
|  | Non-symptomatics | 0.041 ( 0.031 , 0.052 ) | 1.28 ( 1.21 , 1.37 ) | >0.99 | 16.8 ( 22.6 , 13.2 ) |
|  | Positive for both E and N genes | 0.050 ( 0.042 , 0.057 ) | 1.34 ( 1.29 , 1.40 ) | >0.99 | 14.0 ( 16.4 , 12.2 ) |
|  | Positive for both E and N genes or positive only for N gene with CT 35 or less | 0.045 ( 0.038 , 0.052 ) | 1.31 ( 1.26 , 1.36 ) | >0.99 | 15.4 ( 18.1 , 13.4 ) |
| 13 interim | All positives | 0.113 ( 0.057 , 0.172 ) | 1.87 ( 1.40 , 2.45 ) | >0.99 | 6.1 ( 12.3 , 4.0 ) |
|  | Non-symptomatics | 0.200 ( 0.093 , 0.315 ) | 2.75 ( 1.69 , 4.22 ) | >0.99 | 3.5 ( 7.5 , 2.2 ) |
|  | Positive for both E and N genes | 0.085 ( 0.021 , 0.150 ) | 1.63 ( 1.14 , 2.22 ) | >0.99 | 8.1 ( 32.2 , 4.6 ) |
|  | Positive for both E and N genes or positive only for N gene with CT 35 or less | 0.093 ( 0.033 , 0.154 ) | 1.69 ( 1.22 , 2.26 ) | >0.99 | 7.5 ( 21.2 , 4.5 ) |

**Figure 2.**
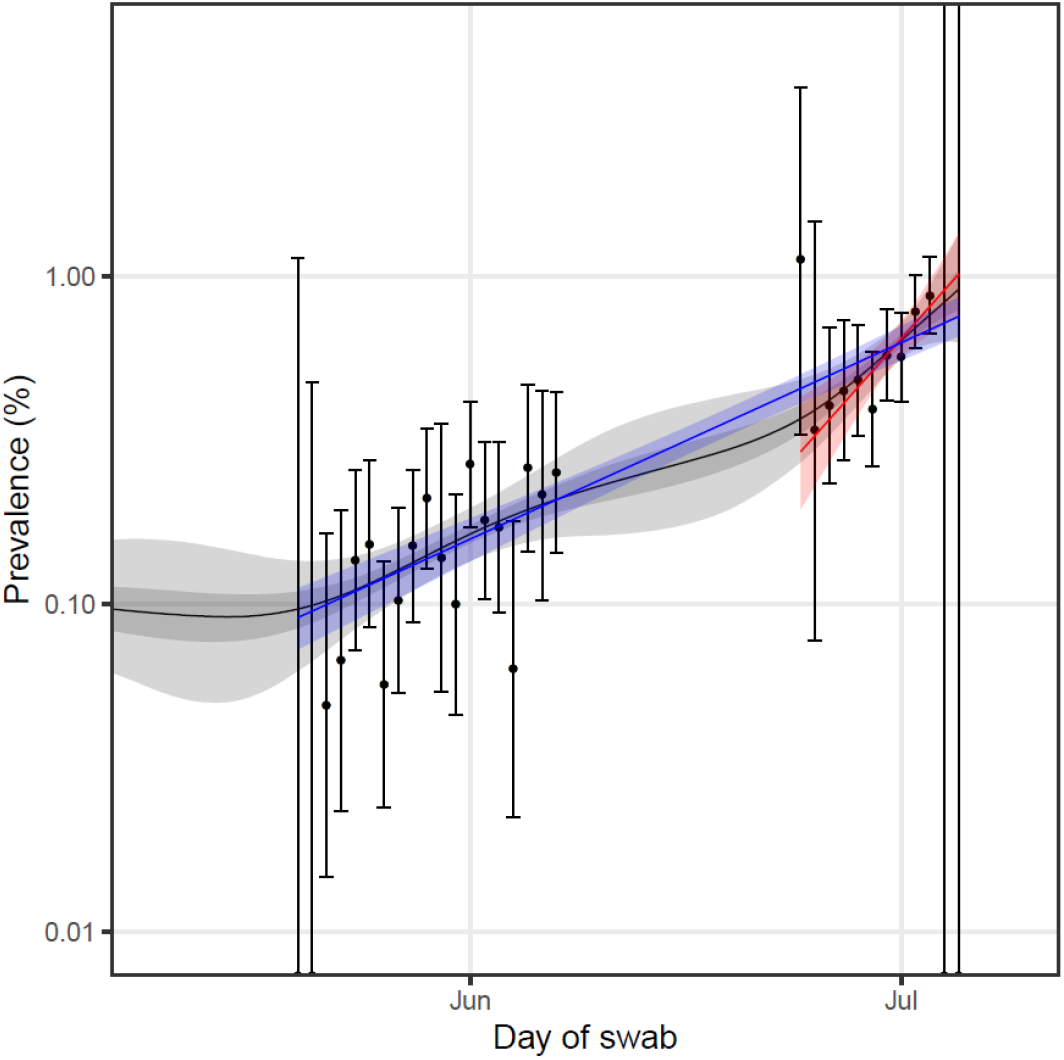
Prevalence of national swab-positivity for England estimated using a P-spline for all twelve completed rounds and round 13 interim with central 50% (dark grey) and 95% (light grey) posterior credible intervals. Best fit exponential model fit to round 12 and round 13 interim of the data (blue) with 95% posterior credible interval. Best fit exponential model fit to round 13 interim only (red) with 95% posterior credible interval. Weighted observations (black dots) and 95% binomial confidence intervals (vertical lines) are also shown. Note that the graph only displays the periods of round 12 and round 13 interim of the study.

Prevalence of swab-positivity was higher among men in round 13 interim at 0.71% (0.58%, 0.88%) compared with 0.47% (0.39%, 0.58%) in women (Table 3a). Women had a reduced odds of 0.69 (0.53, 0.90) of testing positive compared to men after adjustment for core variables (Table 4).

**Table 3a.**
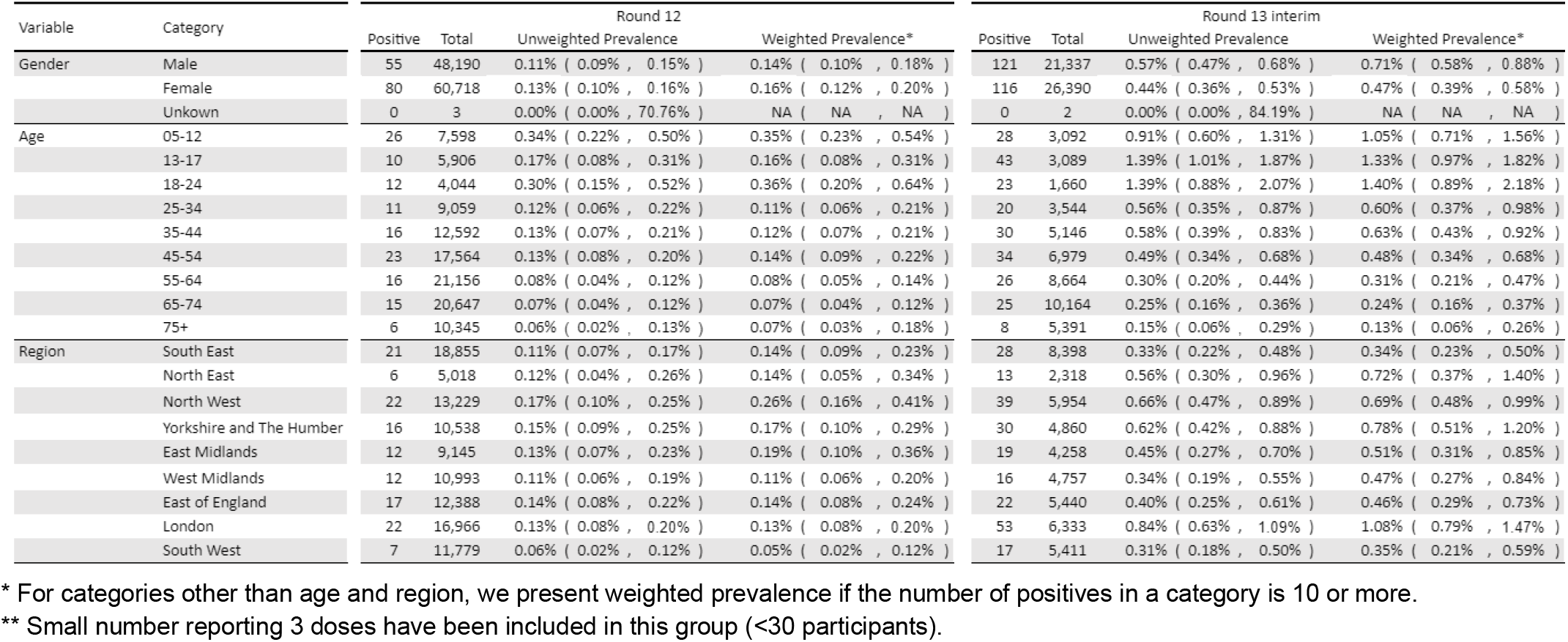
Unweighted and weighted prevalence of swab-positivity for sex, age, and region for round 12 and round 13 interim.

**Table 3b.**
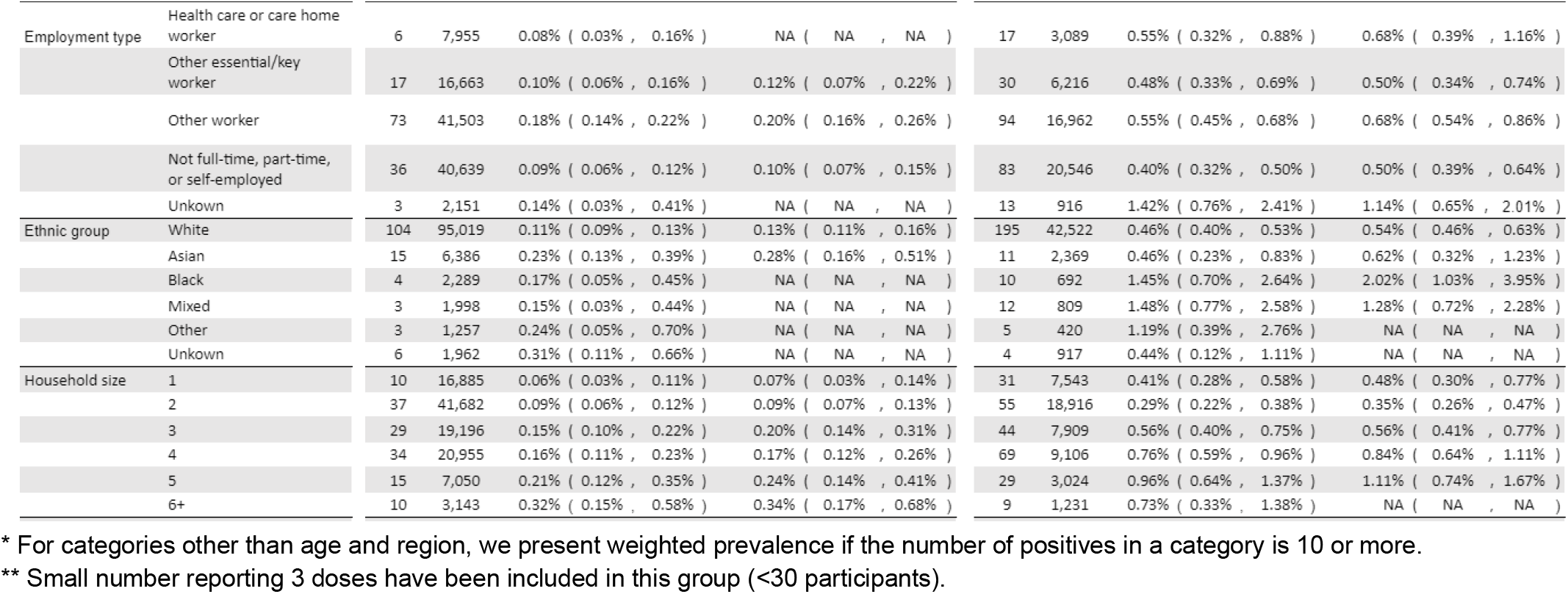
Unweighted and weighted prevalence of swab-positivity for employment type, ethnic group, and household size for round 12 and round 13 interim.

**Table 3c.**
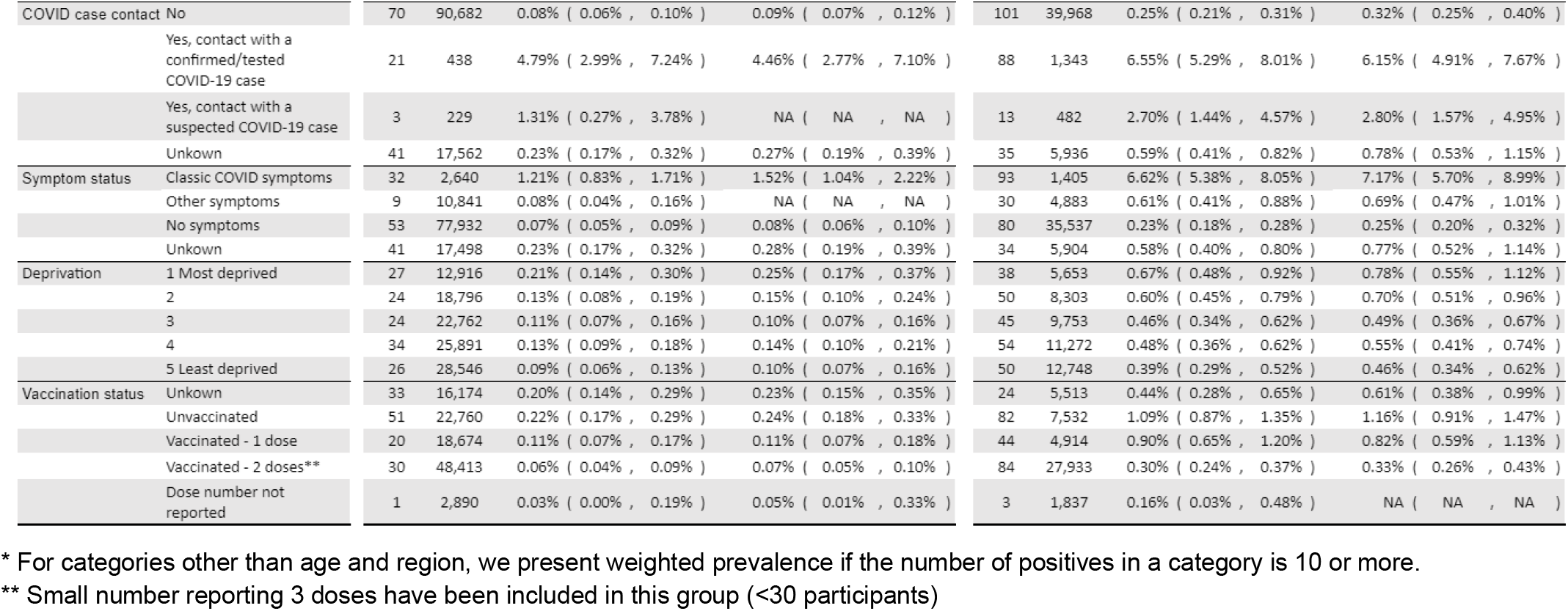
Unweighted and weighted prevalence of swab-positivity for COVID case contact status, symptom status, neighbourhood deprivation and vaccination status for round 12 and round 13 interim.

**Table 4.** Multivariable logistic regression for rounds 12 and round 13 interim.

| Variable | Category | Round 12* | Round 13 interim* |
| --- | --- | --- | --- |
| Sex | Male | Ref | Ref |
|  | Female | 1.34 [0.93,1.92] | 0.69 [0.53,0.90] |
| Key Worker Status | HCW/CHW | 0.37 [0.16,0.87] | 0.98 [0.58,1.66] |
|  | Key worker (other) | 0.55 [0.32,0.94] | 0.83 [0.55,1.27] |
|  | Other worker | Ref | Ref |
|  | Not FT, PT, SE | 0.60 [0.38,0.95] | 0.88 [0.62,1.25] |
| Ethnicity | White | Ref | Ref |
|  | Asian | 1.52 [0.84,2.76] | 0.66 [0.35,1.24] |
|  | Black | 1.26 [0.45,3.55] | 2.11 [1.08,4.12] |
|  | Mixed | 0.95 [0.30,3.05] | 1.39 [0.67,2.87] |
|  | Other | 1.82 [0.57,5.87] | 1.47 [0.54,4.04] |
| Household Size | 1-2 People | Ref | Ref |
|  | 3-5 People | 1.19 [0.75,1.89] | 1.05 [0.74,1.48] |
|  | 6+ People | 1.74 [0.78,3.88] | 0.86 [0.39,1.86] |
| Deprivation Index Quintile | 1 - Most Deprived | 1.94 [1.08,3.51] | 1.34 [0.85,2.10] |
|  | 2 | 1.39 [0.78,2.51] | 1.15 [0.75,1.76] |
|  | 3 | 1.11 [0.62,2.01] | 1.03 [0.68,1.57] |
|  | 4 | 1.55 [0.91,2.63] | 1.16 [0.78,1.72] |
|  | 5 - Least Deprived | Ref | Ref |

\* Odds ratios mutually adjusted for all variables shown.

There have been substantial increases in prevalence in all age groups under the age of 75 years, and especially at younger ages (Table 3a, Figure 3). The highest prevalence was for those aged 13 to 17 years at 1.33% (0.97%, 1.82%) and those aged 18 to 24 years at 1.40% (0.89%, 2.18%). Prevalence in older school-aged children (13 to 17 years) increased eight-fold from 0.16% (0.08%, 0.31%) in round 12.

**Figure 3.**
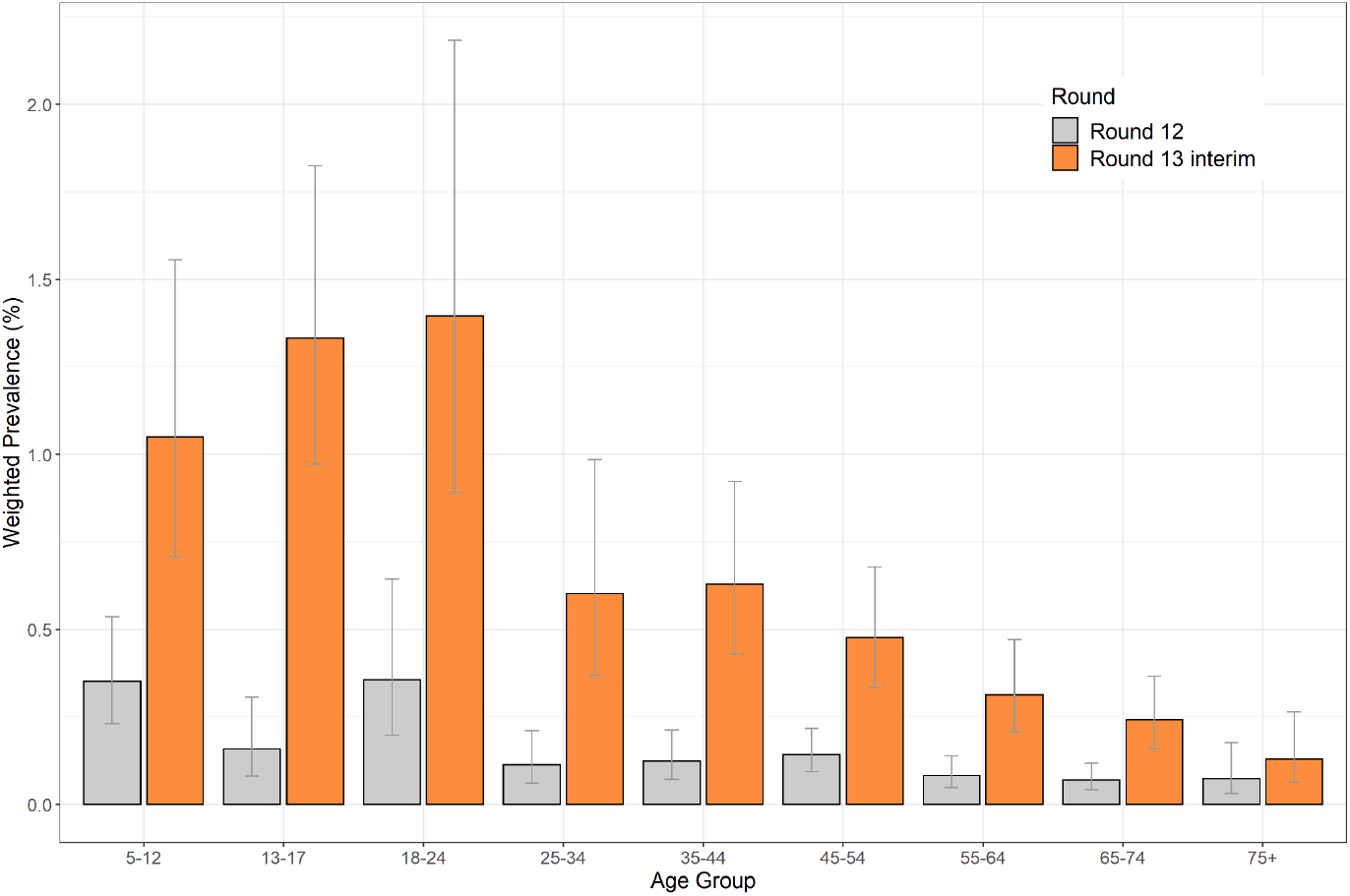
Weighted prevalence of swab-positivity by region for round 12 and round 13 interim. Bars show 95% confidence intervals.

Weighted prevalence of swab-positivity in London increased eight-fold, rising from 0.13% (0.08%, 0.20%) in round 12 to 1.08% (0.79%, 1.47%) in round 13 interim (Table 3a, Figure 4). Using a constant growth rate model at the regional scale, we also detected positive growth for all other regions (Table 5) and observed that the growth rate for the North West was lower than that for London for the period from round 12 to round 13 interim (Table 5).

**Table 5.** Estimates of regional growth rates, doubling times and reproduction numbers for round 12 to round 13 interim.

| Round | Region | Growth rate | R | Probability R>1 | Halving (-) / Doubling (+) time |
| --- | --- | --- | --- | --- | --- |
| 12 and 13 interim | East Midlands | 0.036 ( 0.016 , 0.057 ) | 1.24 ( 1.10 , 1.40 ) | >0.99 | 19.4 ( 44.1 , 12.2 ) |
|  | West Midlands | 0.049 ( 0.027 , 0.073 ) | 1.34 ( 1.18 , 1.53 ) | >0.99 | 14.2 ( 26.1 , 9.4 ) |
|  | East of England | 0.042 ( 0.022 , 0.063 ) | 1.29 ( 1.15 , 1.45 ) | >0.99 | 16.5 ( 31.6 , 10.9 ) |
|  | London | 0.074 ( 0.058 , 0.092 ) | 1.54 ( 1.41 , 1.68 ) | >0.99 | 9.3 ( 12.0 , 7.5 ) |
|  | North West | 0.032 ( 0.017 , 0.046 ) | 1.21 ( 1.11 , 1.32 ) | >0.99 | 22.0 ( 40.0 , 15.0 ) |
|  | North East | 0.061 ( 0.032 , 0.095 ) | 1.43 ( 1.22 , 1.71 ) | >0.99 | 11.3 ( 21.4 , 7.3 ) |
|  | South East | 0.028 ( 0.011 , 0.046 ) | 1.19 ( 1.07 , 1.31 ) | >0.99 | 24.8 ( * , 15.2 ) |
|  | South West | 0.061 ( 0.033 , 0.096 ) | 1.43 ( 1.22 , 1.71 ) | >0.99 | 11.3 ( 20.9 , 7.3 ) |
|  | Yorkshire ** | 0.049 ( 0.031 , 0.068 ) | 1.34 ( 1.21 , 1.49 ) | >0.99 | 14.2 ( 22.3 , 10.2 ) |

\* Doubling/Halving time had an estimated magnitude greater than 50 days and so represented approximately constant prevalence
\*\* Yorkshire and The Humber

**Figure 4.**
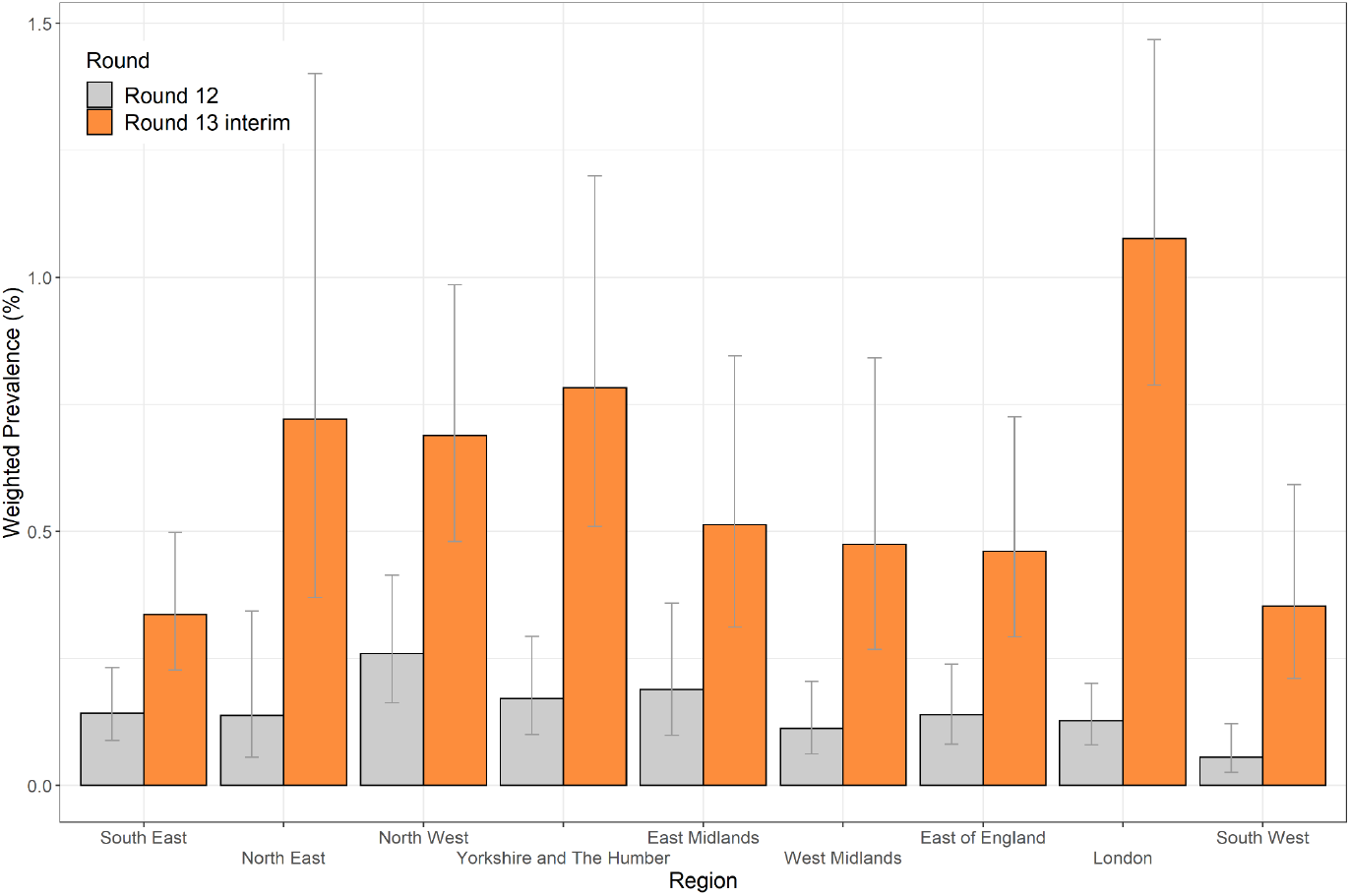
Weighted prevalence of swab-positivity by region for round 12 and round 13 interim. Bars show 95% confidence intervals.

We examined patterns of prevalence according to a number of additional characteristics. Prevalence was higher in people of Black ethnicity at 2.02% (1.03%, 3.95%) compared to people of white ethnicity at 0.54% (0.46%, 0.63%) and this difference, while attenuated, persisted after adjustment for core variables (Table 3b, Table 4). We also observed high rates of swab-positivity among participants who reported having had contact with a confirmed COVID-19 case at 6.15% (4.91%, 7.67%).

We investigated patterns of swab-positivity by self-reported vaccine status. Overall, prevalence was over three times higher in the unvaccinated compared with those reporting two doses of vaccine in both round 12 and round 13 interim (Table 3c). These results allow us to estimate an unadjusted effectiveness for two doses of vaccine against PCR-confirmed swab-positivity of 72.6% (62.8%, 79.6%) for round 13 interim and 72.4% (56.7%, 82.4%) for round 12. In participants aged under 65 years (Table 6): between round 12 and round 13 interim, prevalence increased four- to five-fold in those reporting that they were unvaccinated and those reporting two doses; within round 13 interim prevalence amongst those reporting they were unvaccinated was over three-fold higher at 1.15% (0.92%, 1.43%) compared with those reporting two doses of vaccine at 0.35% (0.26%, 0.45%). In participants aged 65 years and above (Table 6): there was a four-fold increase in prevalence between round 12 and round 13 interim in those who reported two doses of vaccine from 0.06% (0.03%, 0.10%) to 0.24% (0.16%, 0.34%) respectively (Table 6).

**Table 6.** Prevalence of infection for younger and older age groups by self-reported vaccine status.

| Age Group | Vaccination Status | Round 12 |  |  |  | Round 13 interim |  |  |  |
| --- | --- | --- | --- | --- | --- | --- | --- | --- | --- |
|  |  | Negative | Positive | Total | Percentage positive (95% confidence interval) | Negative | Positive | Total | Percentage positive (95% confidence interval) |
| <65 | Status not known | 11107 | 27 | 11134 | 0.24% ( 0.17% , 0.35% ) | 3242 | 19 | 3261 | 0.58% ( 0.35% , 0.91% ) |
|  | Not vaccinated | 21854 | 51 | 21905 | 0.23% ( 0.18% , 0.31% ) | 7027 | 82 | 7109 | 1.15% ( 0.92% , 1.43% ) |
|  | Vaccinated - dose not known | 1176 | 0 | 1176 | 0.00% ( 0.00% , 0.23% ) | 772 | 3 | 775 | 0.39% ( 0.08% , 1.13% ) |
|  | Vaccinated - 1 dose | 18367 | 19 | 18386 | 0.10% ( 0.07% , 0.16% ) | 4844 | 44 | 4888 | 0.90% ( 0.65% , 1.21% ) |
|  | Vaccinated - 2 doses * | 25301 | 17 | 25318 | 0.07% ( 0.04% , 0.11% ) | 16085 | 56 | 16141 | 0.35% ( 0.26% , 0.45% ) |
| 65+ | Status not known | 5034 | 6 | 5040 | 0.12% ( 0.05% , 0.26% ) | 2247 | 5 | 2252 | 0.22% ( 0.07% , 0.52% ) |
|  | Not vaccinated | 855 | 0 | 855 | 0.00% ( 0.00% , 0.32% ) | 423 | 0 | 423 | 0.00% ( 0.00% , 0.87% ) |
|  | Vaccinated - dose not known | 1713 | 1 | 1714 | 0.06% ( 0.01% , 0.33% ) | 1062 | 0 | 1062 | 0.00% ( 0.00% , 0.35% ) |
|  | Vaccinated - 1 dose | 287 | 1 | 288 | 0.35% ( 0.06% , 1.94% ) | 26 | 0 | 26 | 0.00% ( 0.00% , 13.23% ) |
|  | Vaccinated - 2 doses * | 23082 | 13 | 23095 | 0.06% ( 0.03% , 0.10% ) | 11764 | 28 | 11792 | 0.24% ( 0.16% , 0.34% ) |

\* Small number reporting 3 doses have been included in this group (<30 participants).

## Discussion

Initial findings from REACT-1 round 13 based on data from 24 June to 5 July 2021 (round 13 interim) indicate a continuing and accelerating exponential growth in infections since our previous report (round 12, 20 May to 7 June 2021). Prevalence has increased nearly four-fold between round 12 and round 13 interim, giving an overall estimate of approximately 0.6%, equivalent to one in 170. Our estimate of R remains reliably above one with a recent doubling time of around six days (although with wide confidence limits) for the period covered by round 13 interim. This is against a background where Delta variant has now almost completely replaced Alpha variant and is by far the dominant strain of SARS-CoV-2 circulating in England [5].

In the most recent data, risk among men was around 30% higher than that among women, which may reflect different patterns of social mixing in England between men and women. Analysis by age revealed a large increase in infections in 13 to 17 and 18 to 24-year olds, with estimated prevalence among these groups in excess of 1.3%. There were also substantial increases at other ages up to 74 years. We also found large increases in prevalence by region, most notably in London.

Our results indicate that England is now experiencing a substantial third wave of infections, reinforcing other data streams which have been showing a similar signal. Specifically, there has been a rapid increase in the routine reporting of PCR-confirmed cases and positivity (proportion of tested individuals who are positive) [1]. However, the routine data include results of ‘surge’ testing in areas of high prevalence and might overestimate the true increase in the epidemic. Therefore, our results add important context since, being based on a random community sample of named individuals, they should be less affected by changes in testing behaviour than the routine testing. Also, our results show a similar trajectory to those reported by the Office for National Statistics Coronavirus Infection Survey which again uses a community sample (with a household rather than individual-based sample as in REACT-1) [11].

Vaccine status is an important consideration in interpreting our findings. Our crude estimate of vaccine effectiveness against infection after two doses was ∼70% for both rounds 12 and 13 interim. This indicates that there is substantial protection against infection among those who have had two doses of vaccine; however it should be noted that our calculation of effectiveness does not take into account possible differences in behaviour (such as social distancing, mask wearing) among the vaccinated versus unvaccinated population.

The rapidly increasing overall rate of infections in the unvaccinated population is also being reflected in proportional increases among the double-vaccinated population, albeit at much lower levels. But compared with the previous two waves in England, the high efficacy of the vaccine programme against severe disease and death [3] has meant that the increasing infection rate has not, to date, translated into large numbers of people being hospitalised or deaths.

Our study has limitations. While it is based on a random sample of the population, there is variable response among different sectors of the community, such that the results may not be fully representative. We endeavour to overcome any such bias by weighting the sample to be representative of England as a whole. Also, this report is based on interim data from round 13 and therefore does not capture the full picture from the current round which is due to complete data collection on 12 July 2021. As we have seen in previous rounds, underlying trends can change rapidly [13,14]: doubling times averaged over short periods are more volatile than those based on longer periods.

In summary we have documented the continued and accelerating increase of exponential growth of SARS-CoV-2 infections in England from May to early July 2021, as the third wave of infections in England takes hold. We are entering a critical period with a number of important competing processes: continued vaccination rollout to the whole adult population in England, increased natural immunity through infection, reduced social mixing of children during school holidays, increased proportion of mixing occurring outdoors during summer, the intended full opening of hospitality and entertainment and cessation of mandated social distancing and mask wearing. Surveillance programmes are essential during this next phase of the epidemic to provide clear evidence to the government and the public on the levels and trends in prevalence of infections and their relationship to vaccine coverage, hospitalisations, deaths and Long COVID [15].

## Methods

REACT-1 round 13 interim covers the initial period of data collection in round 13, from 24 June to 5 July 2021. As in previous rounds, we approached a random sample of the population in England aged 5 years and above, using the National Health Service (NHS) register of patients. We obtained a self-administered throat and nose swab that was kept refrigerated and then sent by courier for RT-PCR testing, with swabs administered by parent/guardian for children ages 5 to 12 years, and requested that participants complete a short self-administered questionnaire [9]. The sampling procedure in round 12 (20 May to 7 June 2021) and round 13 differed from previous rounds insofar as we selected the random sample to be in proportion to population at lower tier local authority (LTLA) level, whereas previously we aimed for similar numbers of participants in each LTLA. We compared results both unweighted and weighted (to be representative of England as a whole) from round 13 interim to those for round 12. When comparing results with previous rounds (1 to 11) we used weighted data only. The weights take account of variable response according to age, sex, local authority counts, ethnicity and deprivation, and for rounds 1 to 11, for the differences due to the sampling method.

We report trends in prevalence of swab-positivity over time by fitting a smoothed P-spline function to the daily prevalence data (weighted) across all rounds, with knots at 5-day intervals [16]. We applied exponential growth models to estimate the reproduction number R between rounds 12 and 13 interim, and within round 13 interim (although with more uncertainty than the between round estimate due to the limited number of days observed). We also estimated R for differing cut-points of cycle threshold (Ct) values for swab-positivity and among people not reporting symptoms in the week prior to the swab. We estimated vaccine effectiveness from odds ratios in a logistic model.

We analysed the pattern of swab-positivity by age, sex and region, as well as by other socio-demographic variables and reported contact with a COVID-19 case. Statistical analyses were carried out in R [14]. Research ethics approval was obtained from the South Central-Berkshire B Research Ethics Committee (IRAS ID: 283787).

## Data Availability

Supporting data for tables and figures are available either: in the spreadsheet below; or in the inst/extdata directory of the mrc-ide/reactidd github.

https://github.com/mrc-ide/reactidd

## Data availability

Supporting data for tables and figures are available either: in this [spreadsheet](https://docs.google.com/spreadsheets/d/1HzFcneJXlPdbG-FhkMyJT38NKXoncbXTt3vb2BTPW-4/edit#gid=1800680684); or in the inst/extdata directory of this GitHub_R_package.

## Acknowledgements

SR, CAD acknowledge support: MRC Centre for Global Infectious Disease Analysis, National Institute for Health Research (NIHR) Health Protection Research Unit (HPRU), Wellcome Trust (200861/Z/16/Z, 200187/Z/15/Z), and Centres for Disease Control and Prevention (US, U01CK0005-01-02). GC is supported by an NIHR Professorship. HW acknowledges support from an NIHR Senior Investigator Award and the Wellcome Trust (205456/Z/16/Z). PE is Director of the MRC Centre for Environment and Health (MR/L01341X/1, MR/S019669/1). PE acknowledges support from Health Data Research UK (HDR UK); the NIHR Imperial Biomedical Research Centre; NIHR HPRUs in Chemical and Radiation Threats and Hazards, and Environmental Exposures and Health; the British Heart Foundation Centre for Research Excellence at Imperial College London (RE/18/4/34215); and the UK Dementia Research Institute at Imperial (MC_PC_17114). We thank The Huo Family Foundation for their support of our work on COVID-19.

We thank key collaborators on this work – Ipsos MORI: Kelly Beaver, Sam Clemens, Gary Welch, Nicholas Gilby, Kelly Ward, Galini Pantelidou and Kevin Pickering; Institute of Global Health Innovation at Imperial College: Gianluca Fontana, Sutha Satkunarajah, Didi Thompson and Lenny Naar; Molecular Diagnostic Unit, Imperial College London: Prof. Graham Taylor; North West London Pathology and Public Health England for help in calibration of the laboratory analyses; Patient Experience Research Centre at Imperial College and the REACT Public Advisory Panel; NHS Digital for access to the NHS register; and the Department of Health and Social Care for logistic support. SR acknowledges helpful discussion with attendees of meetings of the UK Government Office for Science (GO-Science) Scientific Pandemic Influenza – Modelling (SPI-M) committee.

